# Estimating measures to reduce the transmission of SARS-CoV-2 in Australia to guide a ‘National Plan’ to reopening

**DOI:** 10.1101/2022.12.15.22282869

**Authors:** Gerard E. Ryan, Freya M. Shearer, James M. McCaw, Jodie McVernon, Nick Golding

## Abstract

The availability of COVID-19 vaccines promised a reduction in the severity of disease and relief from the strict public health and social measures (PHSMs) imposed in many countries to limit spread and burden of COVID-19. We were asked to define vaccine coverage thresholds for Australia’s transition to easing restrictions and reopening international borders. Using evidence of vaccine effectiveness against the then-circulating Delta variant, we used a mathematical model to determine coverage targets. The absence of any COVID-19 infections in many sub-national jurisdictions in Australia posed particular methodological challenges. We used a novel metric called Transmission Potential (TP) as a proxy measure of the population-level effective reproduction number. We estimated TP of the Delta variant under a range of PHSMs, test-trace-isolate-quarantine (TTIQ) efficiencies, vaccination coverage thresholds, and age-based vaccine allocation strategies. We found that high coverage across all ages (*≥* 70%) combined with ongoing TTIQ and minimal PHSMs was sufficient to avoid lockdowns. At lesser coverage (*≤* 60%) rapid case escalation risked overwhelming of the health sector or the need to reimpose stricter restrictions. Maintaining low case numbers was most beneficial for health and the economy, and at higher coverage levels (*≥* 80%) further easing of restrictions was deemed possible. These results directly informed easing of COVID-19 restrictions in Australia.

## INTRODUCTION

Since early 2020, rapid dissemination of SARS-CoV-2 and subsequent emergence of variants have resulted in multiple rapidly escalating waves of infection with devastating impacts on health, society and the economy [1]. In contrast with the rest of the world, many island nations in the Western Pacific Region remained relatively COVID free through the first two years of the pandemic as a result of strong border controls to prevent importation, and reactive imposition of social restrictions to constrain community transmission (e.g. [2, 3]). These measures enabled enviable social freedoms and ongoing economic activity, but such disconnection from the international community was not a sustainable strategy over the longer term [4].

Global concerted efforts to accelerate development and licensure of safe and effective vaccines raised hopes that wide scale population immunisation would enable a return to ‘life as normal’ in high burden settings, given demonstrated impacts on infection acquisition and onward spread [5, 6, 7]. For countries that had pursued a low or zero-COVID strategy, ‘living with COVID’ seemed an achievable goal if vaccines could constrain transmission and mitigate disease outcomes sufficiently to avoid overwhelming the health system following SARS-CoV-2 importation, while maintaining near normal societal functioning [8, 9]. However, anticipating the likely impacts of introducing COVID-19 into an environment without established SARS-CoV-2 transmission is difficult [10].

To address this challenge, we use the Transmission Potential (TP) [10] metric to quantify the risk of SARS-CoV-2 transmission in populations based on behavioural, vaccination, and social mixing data. Where the effective reproduction number *R_eff_* is the average number of secondary cases arising from a case in the *infected population*, TP is an estimate of what *R_eff_* would be over the *whole population*, but can be calculated in times of low or no transmission. TP is driven by factors that influence transmission from local cases such as effectiveness of test-trace-isolate-quarantine (TTIQ) case and contact management [11], as well as changes in personal distancing behaviour and mandated constraints on mixing group sizes — collectively termed public health and social measures (PHSMs), and vaccination [10]. TP is not estimated from cases, but instead considers how these influences alter the potential for viral transmission from a baseline *R*_0_ 1. Transmission potential is explained in more detail in the Methods section *Transmission Potential* and in the supplementary section *The relationship of Transmission Potential, R*_0_*, and R_eff_*. Demonstrated reductions in TP achieved by these interventions can be incorporated into future scenario projections, along with anticipated impacts of vaccination on transmission. International evidence of vaccine effectiveness against acquisition and onward spread of the Delta variant was used to estimate the likely overlaid impact of differing levels of vaccine coverage (by age cohort) on population wide TP.

This approach was used in mid-2021 to help inform Australia’s National Plan to transition Australia’s National COVID-19 response[12], by determining target vaccination thresholds for moving between its four phases (table 1), including any ongoing requirement for public health responses and social measures. These methods formed the basis for further collaborative work with the Australian Government Treasury to determine likely economic implications of reopening at alternative thresholds based on the level of adjunct PHSMs needed [13], and constituted part of a broader suite of modelling advice to the Australian Government to inform management of the pandemic and a shift away from strict lockdown measures [14, 15, 16]. From the outset, it was anticipated that reopening targets in populations näıve to COVID-19 would be higher than for those with a history of prior circulation resulting in some degree of established immunity to the virus.

**Table 1.** Phases of the “National Plan to transition Australia’s National COVID-19 Response”[12]. Our modelling analysis focuses on the transition from ‘phase A’ in which strong suppression and no community transmission is the goal, to ‘phase B’ where vaccine coverage is high and SARS-CoV-2 infection is allowed to establish in the population. Scenarios therefore examine the epidemic dynamics and clinical consequences of infections following seeding of an epidemic at different vaccination coverage thresholds achieved through alternative age prioritisation strategies.

| Phase | Description | Activities |
| --- | --- | --- |
| A | Vaccinate, Prepare and Pilot | Continue to strongly suppress the virus for the purpose of minimising community transmission |
| B | Vaccination Transition | Seek to minimise serious illness, hospitalisation and fatality as a result of COVID-19 |
| C | Vaccination Consolidation | As above |
| D | Post-Vaccination | Manage COVID-19 consistent with public health management of other infectious diseases |

## METHODS

### Transmission Potential

Transmission potential (TP) is a measure of the average potential for a virus to spread at the population level [10]. It can be considered as an estimate of the reproduction number, if transmission were widespread (and therefore not concentrated in e.g. one demographic group with non-representative transmission rates). At the baseline, TP is the basic reproduction number of a population *R*_0_, but estimating TP also takes into account the multiplicative effects of transmission reducing influences such as vaccination or behavioural change to produce an estimate of potential transmission specific to a context. The base model consists of three sub-models involving time to detection and isolation of cases, and two types of physical distancing behaviour: we differentiate “macro-distancing” as the reduction in the average rate of non-household contacts (e.g. in response to lockdown-type restrictions), while “micro-distancing” is the reduction in transmission probability per non-household contact (e.g. adherence to social-distancing and hygiene advice or legislation; [10]). Transmission potential will vary among communities by differential average household size and age structures, and can be modulated by changes to behaviour through public health and social measures, time to detection and isolation of cases, and immunity through infection and vaccination. We provide further detail on Transmission Potential in the supplementary section *The relationship of Transmission Potential, R*_0_*, and R_eff_*, while a description of the base model can be found in Golding and colleagues [10]. Below we explain and test how varying and adding components can modulate TP. Whilst an estimate of Transmission Potential under a given scenario of *R*_0_, behaviour, and health system performance can never be a perfect predictor of a reproduction number due to epidemic stochasticity and unaccounted sources of variation and uncertainty, it is a measure that enables prediction of risk in the event of widespread transmission, and has been used as a systematically reported situational assessment metric in Australia since 2020 [17, 18, 19].

### Test, Trace, Isolate, and Quarantine

Test-trace-isolate-quarantine (TTIQ) strategies are key non-pharmaceutical interventions used globally to manage infectious disease outbreaks [20, 21] including frequently during the COVID-19 pandemic [22, 23]. TTIQ operates through isolating cases identified by testing, and tracing and pre-emptive quarantining of their close contacts to prevent further onward transmission. We adapted our TP model to include an explicit effect of reducing the time to case isolation achievable through intensive contact tracing, in addition to the time to case detection effect already included [11]. The empirical distribution of times from symptom onset to case isolation under an ‘optimal’ TTIQ capacity (i.e. with a health system that had enough capacity to rapidly contact trace all cases) was estimated using a limited time series of case data from the state of New South Wales between July 2020 and January 2021, for which dates of isolation were known with a high level of data completeness. This distribution was then calibrated to estimate the distribution of times to isolation in other times and states by assuming improvements in TTIQ are proportional to improvements in times to detection. We characterised a second level of ‘partially’ efficacious TTIQ based on observations from the state of Victoria on 4 August 2020. This date was the then-peak of daily locally-acquired COVID-19 cases in Australia representing a contact-tracing system under resource constraints. These data were used to estimate the effect of ‘partial’ TTIQ on transmission potential to estimate a baseline TP under community transmission. These estimates of ‘optimal’ and ‘partial’ TTIQ correspond to a 54% and 42% reduction in transmission respectively; the full details of this estimation are found in [11].

### Public Health and Social Measures

In the presence of ongoing viral transmission, it is necessary to keep the rate of virus reproduction, *R_eff_*, below or close to 1, as any extended periods where *R_eff_ >* 1 can quickly lead to significant numbers of cases, causing stressors on health systems. So maintenance of *R_eff_* near or below 1 is needed both to contain community transmission in the suppression phase, and to prevent cases from exceeding health sector capacity after re-opening. Regulated or advised risk reduction behaviours and constraints on social mixing described collectively as public health and social measures (PHSMs) are the levers that may be employed to manage *R_eff_* in response to incursions and outbreaks. Behaviours change over time either spontaneously because of heightened concern or complacency, or in response to mandated public health orders invoking various elements of PHSMs.

Here we investigated what level of PHSMs would be required to bring TP to near or below 1 under different scenarios of vaccination coverage. In collaboration with the Australian Government Treasury we defined four ‘bundles’ of PHSM restrictions: baseline, low, medium and high. Each bundle is drawn from a specific time and place in Australia’s pandemic experience, thereby capturing both real-world behavioural responses and the proportional reduction in TP achievable by PHSMs in this context. They therefore represent differing degrees of social constraints for which the impact on TP and economic activity could be defined from historical observations.

Full descriptions of measures included in these bundles are in table S1. These bundles are intended to reflect the plausible behavioural consequences of different levels of stringency of PHSM. As such, the ‘Baseline PHSM’ bundle reflects behaviours during a period with minimal restrictions, and a population aware of the risk of COVID-19 (as opposed to behaviour prior to the pandemic). The other bundles reflect the behavioural effects of increasing levels of stringency. We emphasise that the TPs associated with these PHSM bundles reflect state wide population behaviours (numbers of household contacts and adherence to hygiene advice) estimated at these times. TPs are observed to differ substantially over time and between states, even within similar restrictions [10]. These periods are therefore intended to reflect achievable levels of reduction in TP via PHSMs, rather than inference about the particular impacts of these sets of restrictions.

### Age-structured Vaccination Impact on Transmission

In keeping with WHO guidance [24] and many other countries, Australia’s immunisation program initially prioritised health- and aged-care workers, elderly populations and those at increased risk of transmission and/or severe outcomes [25]. When defining overall target coverage thresholds for the eligible population, an important goal was to consider the distribution of doses received across age categories. While older individuals are more likely to experience severe disease outcomes, young and working adults are expected to make a greater contribution to transmission at the population level. So to explore the effect of age-structured vaccination on transmission, we explored four allocation strategies: oldest first, 40+ years first, all adults, and transmission reducing, described in table 3 (the specific percentages of each age-group vaccinated in these strategies can be found in [16]).These strategies were motivated by being the strategy in place at the time (oldest first), and a series of what were considered the only other practical options. The strategies followed the Australian Therapeutic Goods Administration [26, 27] and Australian Technical Advisory Group on Immunisation [28] advice on which vaccines were administered to which age-groups and the dosing interval between first and second doses 4. We also assume a two week delay from administration until a vaccine takes full effect 4.

**Table 2.** Description of measures implemented under different ‘bundles’ of public health and social measures (PHSMs). Each bundle relates to a specific time and place in Australia’s pandemic experience up to mid-2021 — thereby capturing behavioural responses and the proportional reduction in TP achievable by PHSMs in the Australian context. The proportional reductions in TP observed at each time and place can therefore be related to similar reductions achieved via other combinations of PHSMs (not limited to the ”bundles” in place during the reference period). Similarly, the imposition of any given combination of PHSMs at different times and places may result in variable population responses and thus reductions in TP. More detailed descriptions of the bundles can be found in S1

| PHSM bundle | Description |
| --- | --- |
| Baseline | Minimal density/capacity restrictions and no major outbreaks as in NSW March 2021 |
| Low | More stringent capacity restrictions compared to baseline (*e.g.*, hospitality venues limited to 10 customers per booking), as in NSW 23 August 2020 |
| Medium | Stringent capacity restrictions, group size limits (*e.g.*, fewer than five people outdoors), stay-at-home orders (except work, study, essential purposes), as in NSW 1 July 2021 |
| High | No household visitors, curfew, stay-at-home orders (except essential purposes and permitted work), schools closed (remote learning only), as in VIC 23 August 2020 |

**Table 3.**
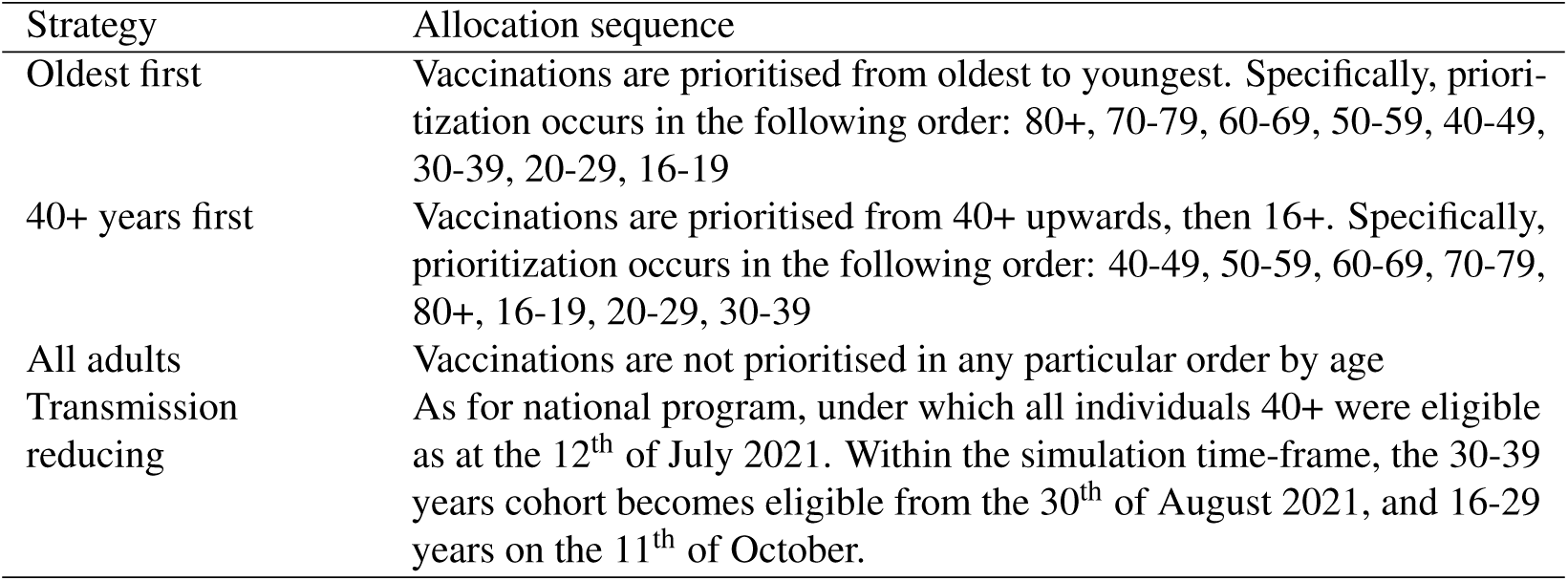
Vaccine allocation strategies and allocation sequence by age-group. The specific modelled percentages of each age-group vaccinated in these strategies can be found in [16].

**Table 4.** Vaccine eligibility, dosing intervals, and assumed delay to efficacy.

| Vaccine | Eligible population | Dosing interval | Delay to full efficacy |
| --- | --- | --- | --- |
| Vaxzevria (ChAdOx1-S) (AZ) [27] | 60+ years [28] | 12 weeks | 2 weeks |
| Comirnaty (BNT162b2) (Pf) [26] | 16+ years | 3 weeks | 2 weeks |

We estimated the percentage reduction in TP that could be expected under different vaccination coverages and distributions by age, vaccine type, and the number of doses received, via static analysis of an age-based transmission matrix, S2, [29]. Age-specific susceptibility and transmissibility estimates [30] are used and transmission rates have been calibrated to a baseline population-wide TP of 3.6. TP will be influenced by spontaneous and imposed changes in physical distancing behaviours, the number of social contacts on average between individuals and the timeliness of TTIQ measures. We use a baseline TP of 3.6 for the Delta variant based on averaged observations from the state of New South Wales in March 2021, a period with minimal social restrictions and no major outbreaks. Although good practice estimation of TP would consider uncertainty in this estimate, the result of this would be to simply shift the scenario estimates up or down the scale. Due to the urgency and nature of the application of this work in providing advice for immediate decision-making, we present the best-estimates only, as used at the time.

For each vaccination scenario, the reduction in transmission by age-group was calculated from the average vaccination efficacy against transmission (accounting for the fractions of each vaccine type and number of doses in that age-group) and the age-group coverage. Proportional reductions in transmission rates for each age combination were then applied to the original ‘unvaccinated’ transmission matrix to construct a ‘vaccinated’ matrix. The dominant eigenvalue, representing the population-wide reproduction number was compared between these pairs of matrices to compute a percentage reduction in TP due to immunisation.

To explore and visualise the effect of different strategies on TP reduction among age categories, we also calculated an age-group specific TP prior to and after vaccination under a given scenario. These ‘by-age’ contributions are calculated for a given homogeneous age-group (whereas other TP calculations use all age groups concurrently S2). Because these age-specific TP calculations exclude interactions with other age-groups, they are not equivalent to the partial contribution of that age group to the overall TP.

### Time in Lockdown

During outbreak suppression in Australia, early and stringent lockdowns were used to bring TP below 1 for the purposes of driving even a handful of local cases from an outbreak to zero, in the context of an optimal TTIQ response. The goal of transitioning to phase B 1 was to minimise the requirement for such stringent PHSMs, restricting their use to meet the explicit objective of prevention of overwhelming the health sector in the face of escalating case loads.

Ongoing application of some degree of social measures through this phase to support vaccine impacts reduces the likelihood for high restrictions and preserves TTIQ effectiveness by keeping case numbers low. TP estimates with and without stringent PHSM can be used to calculate the approximate proportion of time those stringent measures would need to be in place to prevent exceedance of health sector capacity over a defined time frame. This simple static analysis can indicate the plausible societal and economic impacts of the PHSM required to constrain transmission under each scenario and coverage.

Where a vaccination scenario leads to either a *T P*_1_ *>* 1 with one PHSM bundle and *T P*_2_ *<* 1 with a more stringent bundle, the long-term average TP can be maintained at 1, i.e. with daily case counts neither growing nor shrinking over the long term, by alternating between the two PHSM bundle states. Whilst the first PHSM bundle is in place cases will grow, and whilst the more stringent bundle is in place cases will shrink, leading to an oscillation of case counts around some average level S1. This fraction of time under more stringent PHSMs is independent of the sequence or duration of the periods under more stringent restrictions; a strategy of rapid switching on and off of restrictions, or one of alternating long periods with or without restrictions would both lead to long-term maintenance of *TP* = 1, provided the fraction of time in each condition is the same S1.

Switching between more and less stringent PHSMs reflects a strategy that might be used to keep cases below a health sector capacity limit in the event that there is long-term community transmission. With the the necessary simplifying assumption that vaccination coverage is static, where one PHSM bundle leads to growing cases numbers (*T P*_1_ *>* 1), and a second bundle leads to contracting case numbers (*T P*_2_ *<* 1), we can calculate the fraction of time necessary under each bundle of PHSMs S1. However this strategy will not always be either necessary or possible. When cases under both bundles lead to declining case numbers (i.e. *T P*_1_ *<* 1 and *T P*_2_ *<* 1), the fraction is zero as the more stringent PHSM bundle is not needed. Alternatively, where even the more stringent PHSMs still lead to growing daily case numbers (*T P*_2_ *>* 1) no fraction exists, because even the more stringent PHSM bundle could not control transmission.

### Costs of PHSMs

The Australian Government Treasury estimated the direct economic costs of alternative COVID-19 management scenarios explored in this analysis. Estimates included the expected average weekly costs of activity restrictions and lockdowns for each of the bundled levels of restrictions, multiplied by their duration of application over the specified timeframe. These figures were derived by analysing the impact on hours worked across the economy during lockdown periods in 2020, compared with the pre-COVID baseline. They did not include indirect confidence effects, labour market impacts, social, fiscal or health economic costs. For all scenarios it was assumed that case numbers would be constrained by social measures to avoid overwhelming the health system. An objective of maintaining low case numbers in this way was to avoid the significant behavioural changes and related economic impacts that were observed in other country settings where severe and widespread outbreaks occurred in the absence of mixing restrictions. [13].

### Data Analysis

All data analyses for this work were carried out in R [31]. R code to reproduce these analyses are available at https://github.com/aus-covid-modelling/NationalCabinetModelling. This code uses outputs from regular situational assessment work [19, 18] that are conducted using data provided under confidential agreement from the Australian Commonwealth Government (see also statement in Ethics), and that the authors are not authorised to make available. Code to create figures 3 and S1 can be found in [32].

## RESULTS

From an *R*_0_ of 8 for the Delta variant, with baseline PHSMs and partial TTIQ in place TP is reduced to 3.6, which serves as a baseline to which other interventions are added 1. The effects of vaccination and more stringent PHSMs on TP are mathematically multiplicative, so the results displayed in figure 1 use a logarithmic y-axis in order to easily see the relative magnitude of each intervention. The results demonstrate that as vaccination coverage increases, less stringent PHSMs are required to bring TP below 1 and thus control epidemic activity (figure 1). Maintaining a rapid and highly effective TTIQ response capacity is critical for ongoing epidemic control. Should TTIQ responses become only partially effective due to high caseloads, high PHSM would be needed to curb transmission at the 50% and 60% coverage thresholds, whilst low PHSMs may be sufficient for control at 80% coverage (figure 1). More optimistically, the combination of 70% vaccine coverage and ongoing low PHSMs would likely be sufficient for control, if optimal TTIQ can be maintained (figure 1). Note that compliance with imposed measures will vary their effectiveness between populations and time-points. This uncertainty is conceptually represented by the upper and lower bounds of each ‘box’ for each set of restrictions in figure 1.

**Figure 1.**
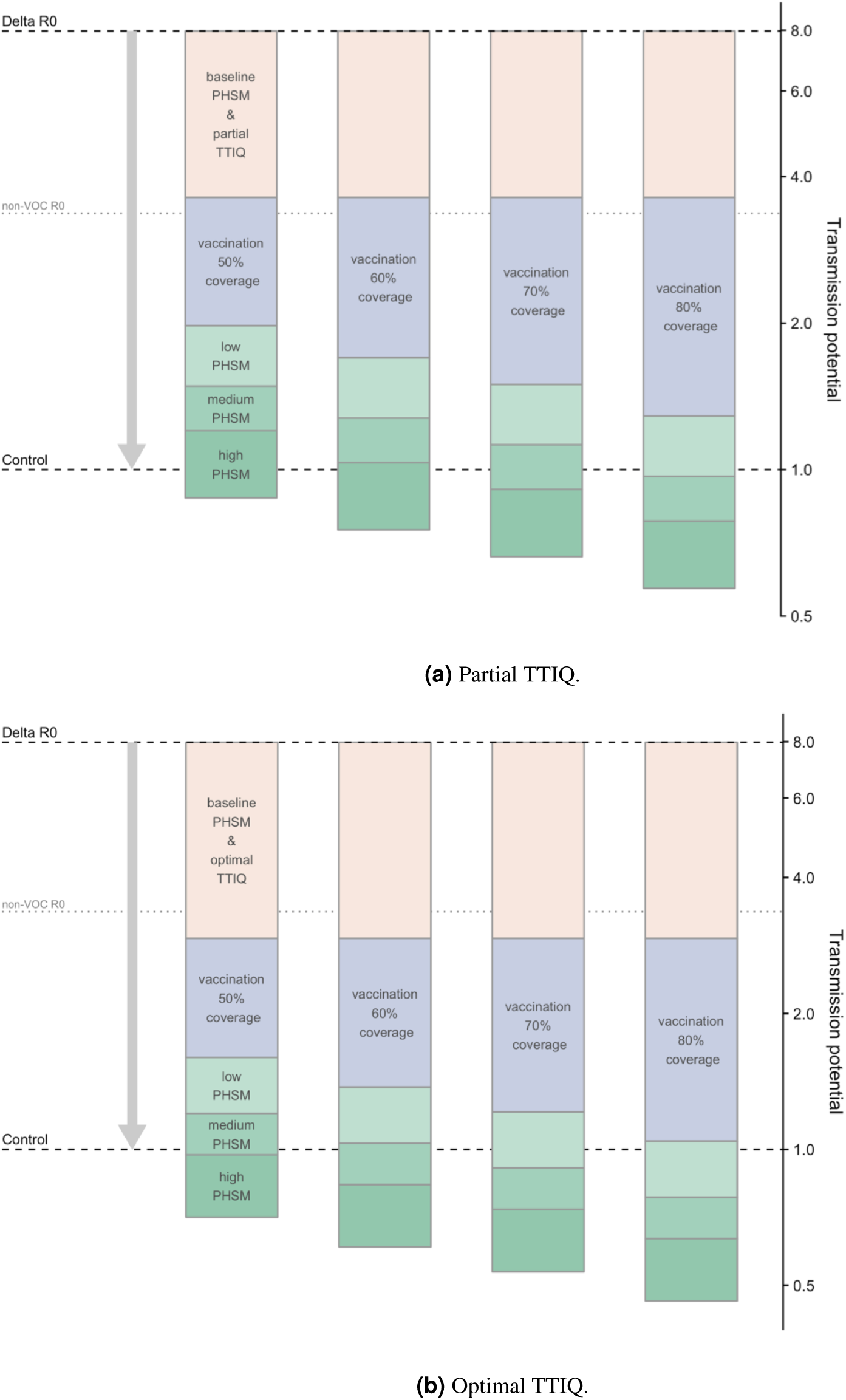
Combined effects of vaccination and PHSM scenarios on COVID-19 transmission potential under the ‘Transmission reducing’ vaccination scenario assuming only partial (a) or optimal (b) TTIQ effectiveness, due to high caseloads. (NB the logarithmic scale of the y-axis enables comparison of the multiplicative effect sizes of these measures without depending on the order of in which they are plotted.)

The choice of age-structured vaccine allocation strategy has a slight effect on Transmission Potential, though this varies with level of vaccine coverage (table 5). The contribution varies considerably by age-group due to differential mixing rates (figure 2, and figures S3, S4, and S5), and while a transmission reducing allocation strategy tends to best reduce TP as intended the effect is slight, and the advantage depends on the overall coverage level of vaccination. Vaccinating the 40+ years first tended to perform worst at lower levels of coverage, requiring the largest proportion of time under strict PHSMs, however this became unimportant at higher levels of vaccination.

**Figure 2.**
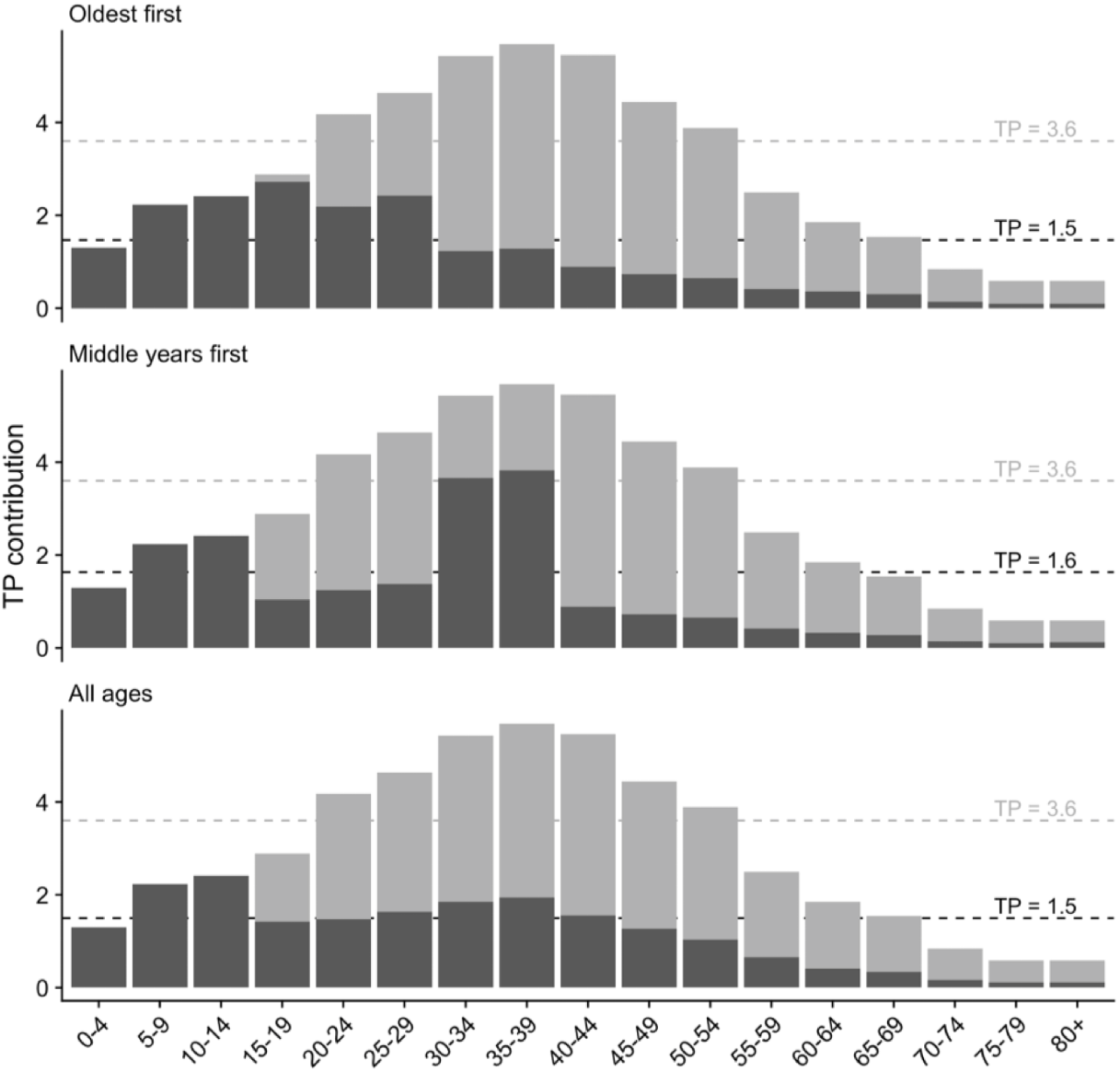
Impact of the four different vaccine allocation strategies on TP by age category, resulting in the overall TP achieved by 70% age eligible population coverage. Dark grey represents TP contribution after vaccination, and light grey in the absence of vaccination. Other coverage levels (50, 60 and 80% are in figures S2a-c). Dashed lines correspond to whole population differences in TP with age-group interactions included.)

**Table 5.**
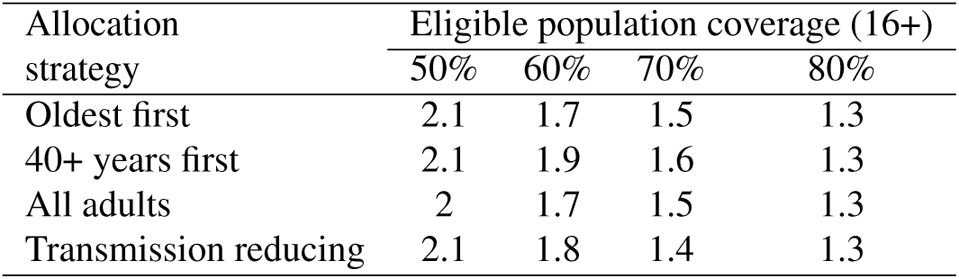
Scaled values of Delta variant transmission potential (TP) for 50%, 60%, 70% and 80% population vaccination coverage for each allocation strategy, with vaccines delivered per 4. We use a baseline TP of 3.6, which corresponds to TP under baseline PHSMs and partial TTIQ.

| Allocation strategy | Eligible population coverage (16+) |  |  |  |
| --- | --- | --- | --- | --- |
|  | 50% | 60% | 70% | 80% |
| Oldest first | 2.1 | 1.7 | 1.5 | 1.3 |
| 40+ years first | 2.1 | 1.9 | 1.6 | 1.3 |
| All adults | 2 | 1.7 | 1.5 | 1.3 |
| Transmission reducing | 2.1 | 1.8 | 1.4 | 1.3 |

Tables 6 and 7 compare the proportion of time that would need to be spent with high PHSM on top of ongoing light restrictions to maintain case counts at some level, by vaccine coverage and allocation strategy. We assume periodic switching between low PHSM and high PHSM over a long period with the same vaccination coverage. With long-term coverage held at 50%, 60%, or 70%, high PHSM would be needed for significant fractions of time (18-89%) if caseloads escalate, leading to ‘partial’ TTIQ effectiveness. For the ‘optimal’ TTIQ scenario and an achieved adult population coverage of 70%, high PHSM would be needed rarely if at all.

**Table 6.** Percentage of time high PHSM would need to be in place for long-term control, with reversion to low PHSM at other times, for 50%, 60%, 70% and 80% population coverage achieved under the three age-based allocation strategies. These scenarios assume partial TTIQ effectiveness, under high caseloads. Standard age (60+) and dosing interval (12 weeks) recommendations are assumed for AZ vaccine.

| Allocation strategy | Eligible population coverage (16+) |  |  |  |
| --- | --- | --- | --- | --- |
|  | 50% | 60% | 70% | 80% |
| Oldest first | 82% | 49% | 18% | 0% |
| 40+ years first | 89% | 67% | 39% | 2% |
| All adults | 75% | 46% | 22% | 0% |
| Transmission reducing | 87% | 52% | 10% | 0% |

**Table 7.**
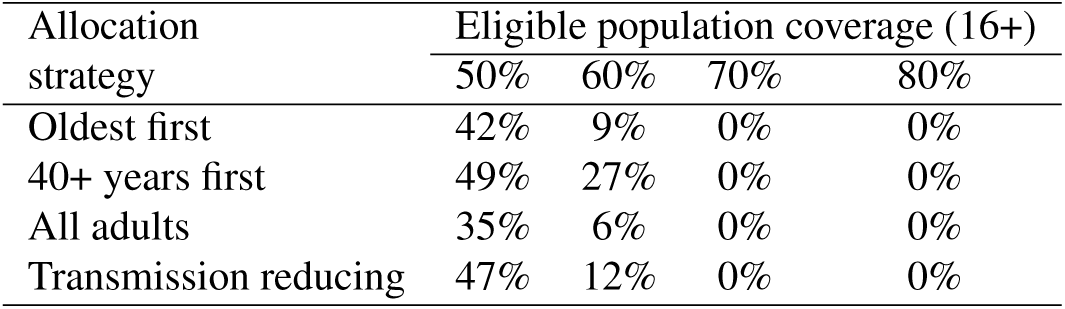
Percentage of time high PHSM would need to be in place for long-term control, with reversion to low PHSM at other times, for 50%, 60%, 70% and 80% population coverage achieved under the three age-based allocation strategies. These scenarios assume optimal TTIQ effectiveness, under high caseloads. Standard age (60+) and dosing interval (12 weeks) recommendations are assumed for AZ vaccine.

| Allocation strategy | Eligible population coverage (16+) |  |  |  |
| --- | --- | --- | --- | --- |
|  | 50% | 60% | 70% | 80% |
| Oldest first | 42% | 9% | 0% | 0% |
| 40+ years first | 49% | 27% | 0% | 0% |
| All adults | 35% | 6% | 0% | 0% |
| Transmission reducing | 47% | 12% | 0% | 0% |

Figure 3 represents the proportion of time spent under each of the restriction stringency settings for different levels of vaccine coverage and intensity of case finding and management strategies, with corresponding costs of these restrictions as estimated by Treasury shown on the right of the figure [13]. These combined outputs demonstrate the substantive cost savings associated with avoidance of lockdown and provided additional justification for delaying reopening until achieving 70% threshold vaccine coverage. The overlay of some degree of social measures at this threshold supported transmission reduction and helped to maintain the effectiveness of an active case finding strategy focused on minimising health impacts. At 80% coverage these restrictions could be eased without any envisaged lockdown requirement.

**Figure 3.**
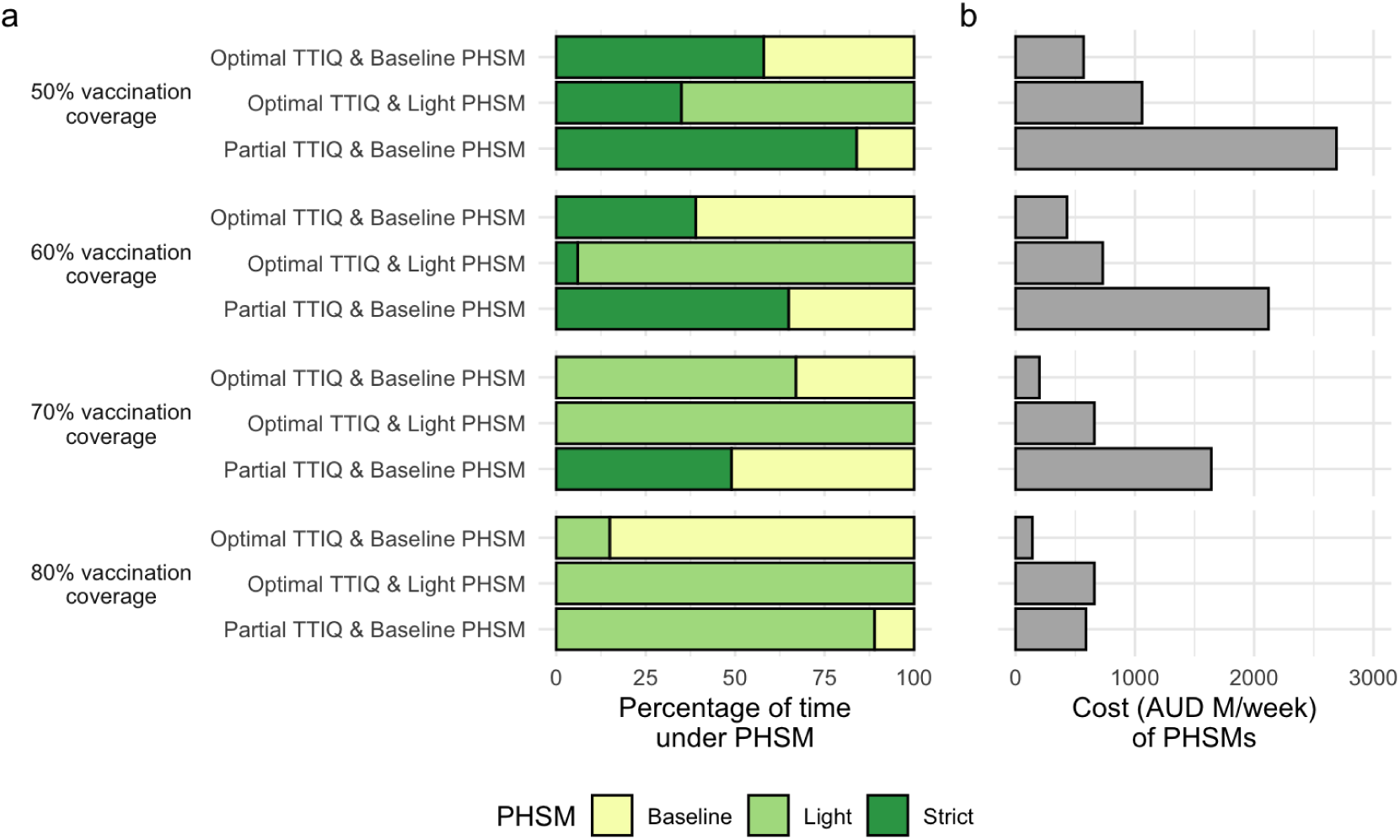
Percentage of time necessary under PHSMs to constrain outbreaks (a) and cost (in millions of Australian dollars per week) of the least expensive PHSM bundle able to constrain outbreaks (b) under each combination of TTIQ effectiveness and ongoing PHSMs, at 50, 60, 70, and 80% vaccination coverage of the 16+ adult population, and an ”All adults” vaccine allocation scenario. Data in (a) from S2, S3, S4, and S5, and (b) from [13]

## DISCUSSION

This work formed the basis of a significant contribution to easing of COVID-19 restrictions in Australia in late 2021 by providing a structured framework to explore the effect of management decisions on transmission of SARS-CoV-2 in Australia based on contemporary evidence [13, 14, 15, 16]. Specifically, it covers the pandemic period up to mid-late 2021, during which time the Delta variant became established in several eastern Australian states resulting in imposition of stringent lockdowns to manage transmission and clinical burden, and closure of interstate borders. For those states, our recommendation of a target vaccination level of at least 70% 1 was a pathway out of restrictions and a way to reconnect with ‘COVID-zero’ jurisdictions. In the other states, high vaccine coverage provided confidence that COVID-19 impacts could be mitigated sufficiently to avoid substantive health system and social disruption, and at 80% coverage only minimal social and behavioural measures would be required to support vaccination 1.

The scenarios in this study representing a single national COVID-19 epidemic were clearly (and deliberately) artificial and served to inform high level policy strategy. Beyond defining threshold vaccine targets, they highlighted the importance of a combination of timely public health responses (TTIQ) and ongoing social and behavioural measures (PHSMs) to constrain SARS-CoV-2 transmission. It was recognised that at high caseloads, maintenance of optimal TTIQ was likely infeasible. In such instances, jurisdictions retained the flexibility to strengthen PHSMs generally or locally (as envisaged in the National Plan [12]) to regain local epidemic control 3. Although our work found little impact of age-structured vaccination on transmission outcomes 5, it is likely that such structure does have an effect on other outcomes like severe disease or death. A further limitation of the need for high-level policy advice was a reliance on best-estimates rather than considering the broader suite of uncertainty around these parameters. Due to the dynamic pandemic situation, uncertainty was instead managed by ongoing surveillance, with the ultimate required intensity and duration of measures informed by ongoing situational assessment of transmission and its related health impacts [19].

In August 2021 when these analyses were first reported, evidence of vaccine efficacy against acquisition and onward spread of infection had raised hopes that equitable vaccine distribution could substantially limit global transmission and burden of COVID-19 disease [33]. However, studies undertaken by multiple modelling groups supporting decision making to ease UK lockdowns during the Delta era similarly cautioned against the lifting of all restrictions (‘freedom day’) following achievement of vaccine targets, given imperfect vaccine protection and potential for resurgence [34]. In support of this position, a rapid rebound of Delta infections following reopening in the Netherlands had necessitated re-imposition of social measures within only weeks of easing. At that time, global concern about driving emergence of further variants remained high and strong suppression strategies were favoured by the World Health Organisation [35].

Our findings were aligned with and benchmarked against the UK modelling reports, including the recommendation for ongoing social constraints, but were novel in two main aspects. For the majority of Australian jurisdictions, relaxation of border restrictions would allow importation of infections into ‘COVID-zero’ settings, requiring high confidence in model recommendations. The use of transmission potential as a novel metric enabled anticipation of case loads in settings where there were presently none [10]. Given ongoing global discussions about competing health and economic impacts of COVID-19 and social measures for its control [36], the accompanying Treasury analyses were highly influential in whole of government decision making. We are unaware of similar publicly available estimates of the costs of PHSMs under different levels of vaccine coverage from other country settings.

The model assumed fixed efficacy of vaccine protection against transmission and disease at a single time point and did not incorporate waning immunity. At the time of analysis, longevity of population experience with COVID-19 vaccines was limited and evidence of the rate of loss of protection was sparse. Israel was the first country to approve booster doses on July 30 2021, initially for individuals aged 60 and above. This recommendation was prompted by observation of breakthrough infections following primary vaccination consistent with waning immunity [37]. Booster requirements were then a contentious issue, with many arguing that broad provision of a third vaccine dose was unnecessary in highly immunised populations. There were particular concerns about impacts on supply for global vaccine equity, given persistently low primary vaccine coverage and access in many low and middle income countries [38].

Note that the visualisations and metrics presented in this paper were used to evaluate the general viability of different suites of measures, under a very uncertain future epidemiological situation and period of time. These metrics were not sufficient to calculate the likely morbidity and mortality outcomes of COVID-19 under specific rollout strategies and changes given the dynamic nature of vaccination and transmission. Related work extended our initial findings on transmission potential into an agent based model framework to estimate those impacts and is reported elsewhere [39].

It was further recognised that the national COVID-19 epidemic had been, and would continue to be, a ‘fire’ fought on multiple fronts across Australia’s geographically distributed population, largely concentrated in coastal urban cities. We recommended that particular attention be paid to groups in whom socioeconomic, cultural and other determinants were anticipated to result in higher transmission and/or disease outcomes. In addition, achievements of vaccination targets at small area level was critical to ensure equity of program impact, as ongoing outbreaks in under-vaccinated populations were considered likely, and would need to be supported by optimisation of localised public health responses. These issues were the focus of subsequent work [11, 39].

In reality, the applicability of these defined thresholds to reopening goals was made redundant by importation and rapid transmission of the Omicron SARS-CoV-2 variant in December 2021. Dissemination of this and subsequent immune escape variants has led to global reconsideration of the role of vaccination in the control of COVID-19. Moreover, vaccine protection against severe disease appears more robust and sustained than that against transmission, particularly in the context of waning post-immunisation neutralising antibody titres [40]. This emerging understanding has reoriented strategic vaccine use towards promoting population resilience against severe disease outcomes, rather than transmission reduction. Subsequent work focuses on the implications of variant emergence for deployment of vaccines and other control measures in the era of Omicron and beyond.

## Data Availability

Code to reproduce these analyses are available at, https://github.com/aus-covid-modelling/NationalCabinetModelling; Code to create figures 3 and S1 can be found in https://doi.org/10.5281/zenodo.7117840. 
This code uses outputs from regular situational assessment work described in https://www.medrxiv.org/content/10.1101/2021.11.28.21264509v1 that are conducted using data provided under confidential agreement from the Australian Commonwealth Government.

https://doi.org/10.5281/zenodo.7117840

## ETHICS

The study was undertaken as urgent public health action to support Australia’s COVID-19 pandemic response. The study used data from the Australian National Notifiable Disease Surveillance System (NNDSS) provided to the Australian Government Department of Health under the National Health Security Agreement for the purposes of national communicable disease surveillance. Data from the NNDSS were supplied after de-identification to the investigator team for the purposes of provision of epidemiological advice to government. Contractual obligations established strict data protection protocols agreed between the University of Melbourne and sub-contractors and the Australian Government Department of Health, with oversight and approval for use in supporting Australia’s pandemic response and for publication provided by the data custodians represented by the Communicable Diseases Network of Australia. The ethics of the use of these data for these purposes, including publication, was agreed by the Department of Health with the Communicable Diseases Network of Australia.

## ACKNOWLEDGEMENTS

This work forms part of a wider body of work around supporting Australia’s plan to reopen from COVID-19 restrictions (often known in media as ’the Doherty modelling’). We thank our many colleagues who, while largely locked in their homes, contributed to this large undertaking with unfailing good grace and humour.

We also thank many colleagues at Treasury who helped advise and focus this work, in particular including those contributing to [13].

Thanks to David Price for helpful comments on the manuscript.

This work was directly funded by the Australian Government Department of Health Office of Health Protection. Additional support was provided by the National Health and Medical Research Council of Australia through its Centres of Research Excellence (SPECTRUM, GNT1170960) and Investigator Grant Schemes (JMcV Principal Research Fellowship, GNT1117140; FMS Emerging Leader Fellowship, 2021/GNT2010051).

This research was supported by The University of Melbourne’s Research Computing Services and the Petascale Campus Initiative.

## SUPPLEMENTARY MATERIALS

### The relationship of Transmission Potential, *R*_0_, and *R_eff_*

*R*_0_ is the number of secondary infections from an infection in a given, fully susceptible population under standard mixing behaviours.

*R_eff_* is the average number of secondary infections from an infection in the *infected population* at a given point in time. In a fully susceptible population this is likely to approximate *R*_0_ but will vary due to levels of immunity in the population, changes in behaviour, and the specific infected population. *R_eff_* is a real, occurring quantity which is can be estimated from records of infections.

Transmission potential, *TP*, is an estimate of the expected *R_eff_* over *whole population*. *TP* is a theoretical quantity used to understand transmission risk over a broader population, in particular in times when transmission is low or zero [10]. Where transmission is widespread, *R_eff_* may be similar to *TP*, and where a population is fully susceptible, and no measures are in place to reduce transmission, *TP* will equate to *R*_0_ (e.g. in main text figure 1, without PHSMs, TTIQ, or vaccination).

### Fraction of time in lockdown

Where a vaccination scenario leads to either a *T P*_1_ *>* 1 with one PHSM bundle and *T P*_2_ *<* 1 with a more stringent bundle, the long-term average TP can be maintained at 1, i.e. with daily case counts neither growing nor shrinking over the long term, by alternating between the two PHSM bundle states. Whilst the first PHSM bundle is in place cases will grow, and whilst the more stringent bundle is in place cases will shrink, leading to an oscillation of case counts around some average level S1. This reflects a strategy that might be used to keep cases below a health sector capacity limit in the event that there is long-term community transmission and under the necessary simplifying assumption that vaccination coverage is static.

Where *T P*_1_*>* 1 and *T P*_2_*<* 1, we can calculate the fraction of time spent under more stringent PHSMs

as:

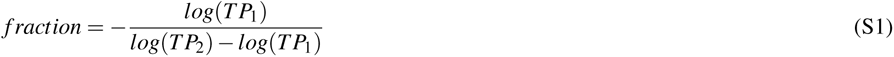

Where *T P*_1_ *<* 1 the fraction is zero, and as the more stringent PHSM bundle is not needed. Where *T P*_2_ *>* 1 no fraction exists, because even the more stringent PHSM bundle could not control transmission.

Note that this fraction of time under more stringent PHSMs is independent of the sequence or duration of the periods under more stringent restrictions; a strategy of rapid switching on and off of restrictions, or one of alternating long periods with or without restrictions would both lead to long-term maintenance of *TP* = 1, provided the fraction of time in each condition is the same and that daily case numbers are either growing or contracting exponentially (i.e., no significant susceptible depletion alters the growth rate), e.g. S1. However for practical reasons, a rapid switching between states is unlikely to be used. The overall number of cases (as opposed to the long-term average TP) will however be dependant on the rate of switching, but it is assumed in all calculations in this paper that switching is sufficient to maintain total case numbers below a level which would result in overwhelming of the public healthcare system.

**Figure S1.**
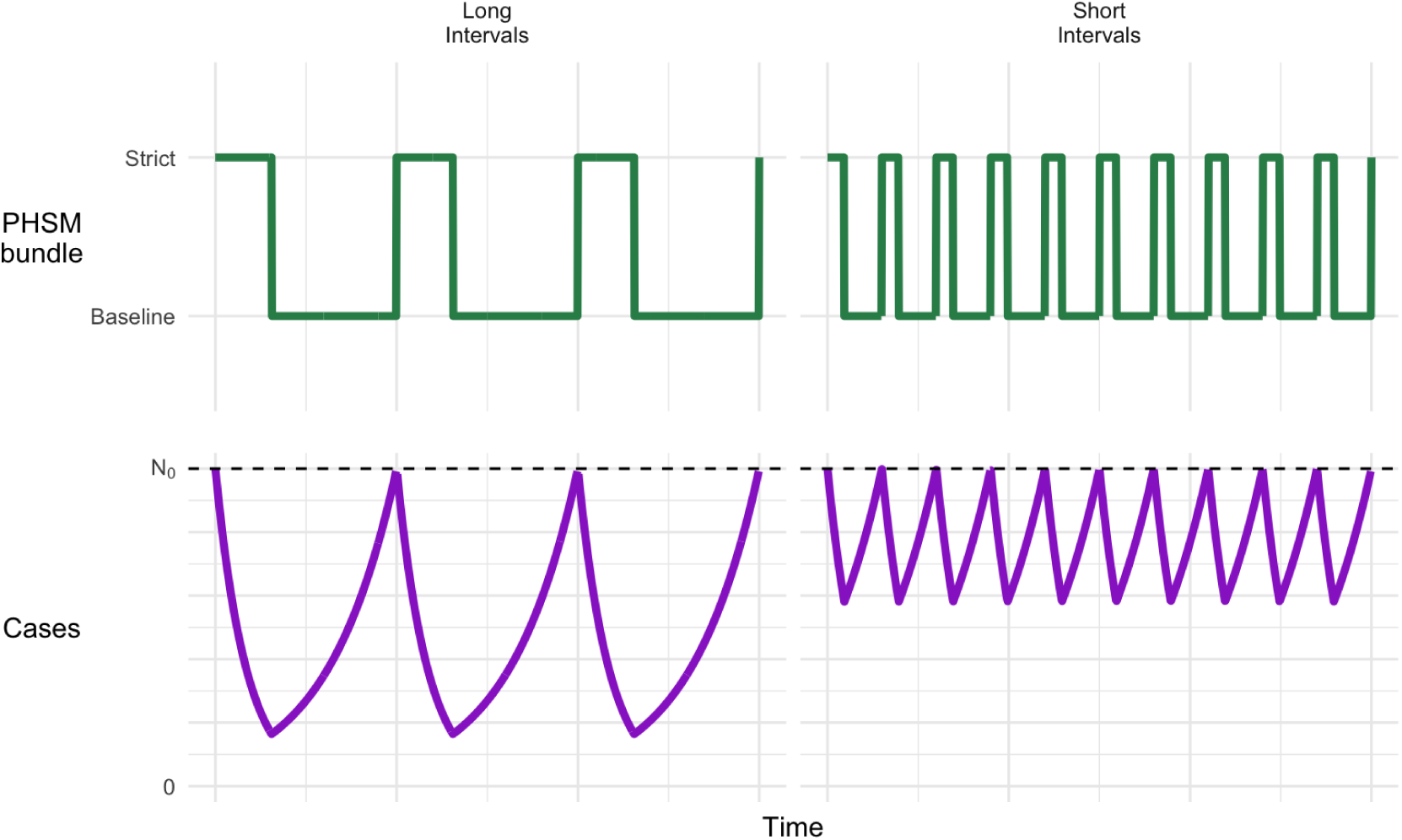
Example of how daily cases may fluctuate over time in response to stringency of PHSMs using 80% vaccination coverage, partial TTIQ and reversion to baseline PHSMs, with either long or short intervals between switching PHSMs. In this example, to maintain a stable TP *≤* 1 over time requires strict PHSMs for *≥* 31% of the time 3, S2. Starting with strict PHSMs as here results in cases fluctuating below the initial daily case number (*N*_0_, dotted line). The length of intervals under respective PHSMs (left vs. right) will not alter the overall average TP (though may affect total case numbers).

### Transmission matrix

Population mixing within and between age groups is configured based on widely accepted social contact matrices published by [29]. It has been expanded to include an 80+ age class (assumed to have the same mixing rates as 75-79 years) S2. Age-specific susceptibility and transmissibility estimates from [30] are used and transmission rates have been calibrated to our baseline population-wide TP of 3.6.

The greatest mixing intensities are anticipated between individuals aged from 15-24 years, remaining high through adults of working age. While intense school-based mixing is anticipated between children aged 5-14, the transmission matrix accounts for the relatively low observed infectiousness of this age group, associated with a high proportion of asymptomatic infections.

**Figure S2.**
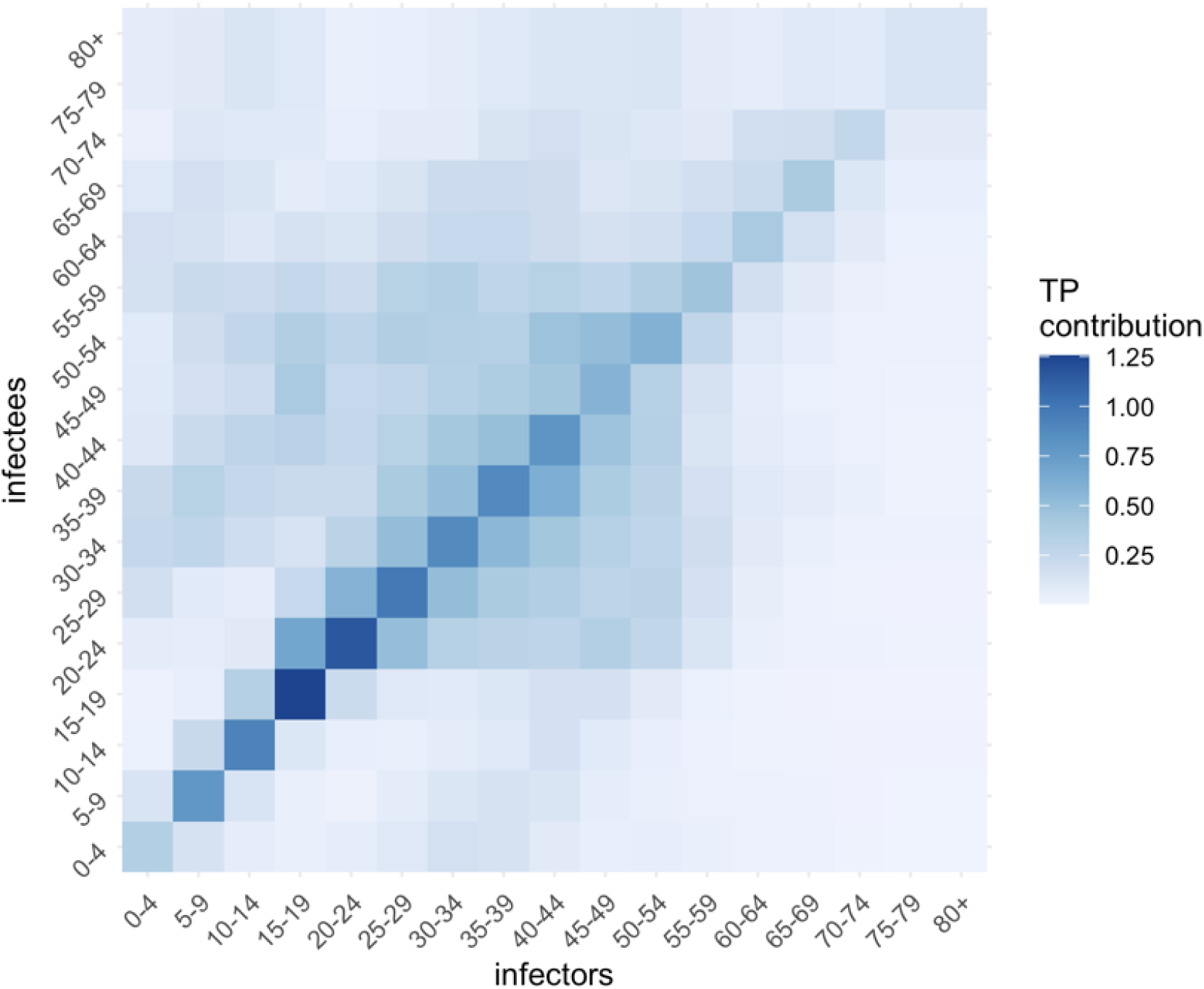
Age-based transmission matrix derived from [29].

**Table S1.** Detailed description of measures implemented under PHSM ‘bundles’.

|  | High PHSM | Medium PHSM | Low PHSM | Baseline PHSM |
| --- | --- | --- | --- | --- |
| Reference period | VIC 23 August 2020 | NSW 1 July 2021 | NSW 23 August 2020 | NSW March 2021 |
| Stay at home orders | Stay-at-home except essential purposes | Stay-at-home except for work, study and essential purposes | No stay-at-home orders | No stay-at-home orders |
| Density restrictions | 4 m<sup>2</sup> rule | 2 m<sup>2</sup> rule | 2 m<sup>2</sup> rule | 2 m<sup>2</sup> rule |
| Retail trade | Non-essential retailers and venues closed to public. Take away and home delivery only | Increased retail activity, subject to density restrictions. Seated dining for small groups at cafes/restaurants | Social distancing rules apply. Larger groups allowed. | Social distancing rules apply |
| Work | Only workplaces categorised as permitted work allowed to operate on-site and subject to restrictions | Work from home if possible, capacity limits and restrictions on office space apply | Return to work, but social distancing and capacity restrictions on office space apply | 1.5 m<sup>2</sup> rule |
| Schools and childcare | Closed – remote learning only | Closed or graduated return | Open | Open |
| Capacity restrictions | No gatherings - Non-essential venues etc closed. | Indoor venues closed. Capacity limits restricted to small groups outdoors | Recreational activities allowed and venues open but social distancing and capacity limits apply | Large sporting venues to operate at 70 per cent capacity |
| Travel restrictions | Essential movements only within 5 or 10 km radius. No intra- or inter-state travel | Non-essential travel limited. No intra- or inter-state travel | No travel restrictions. Interstate travel allowed | No travel restrictions. Interstate travel allowed |
| Other | Curfew. No household visitors and 2-person limit on exercise | 5 visitors to household and limited outdoor gatherings e.g., 10 people | Requirements for record keeping, COVID-safe plans etc. |  |

**Figure S3.**
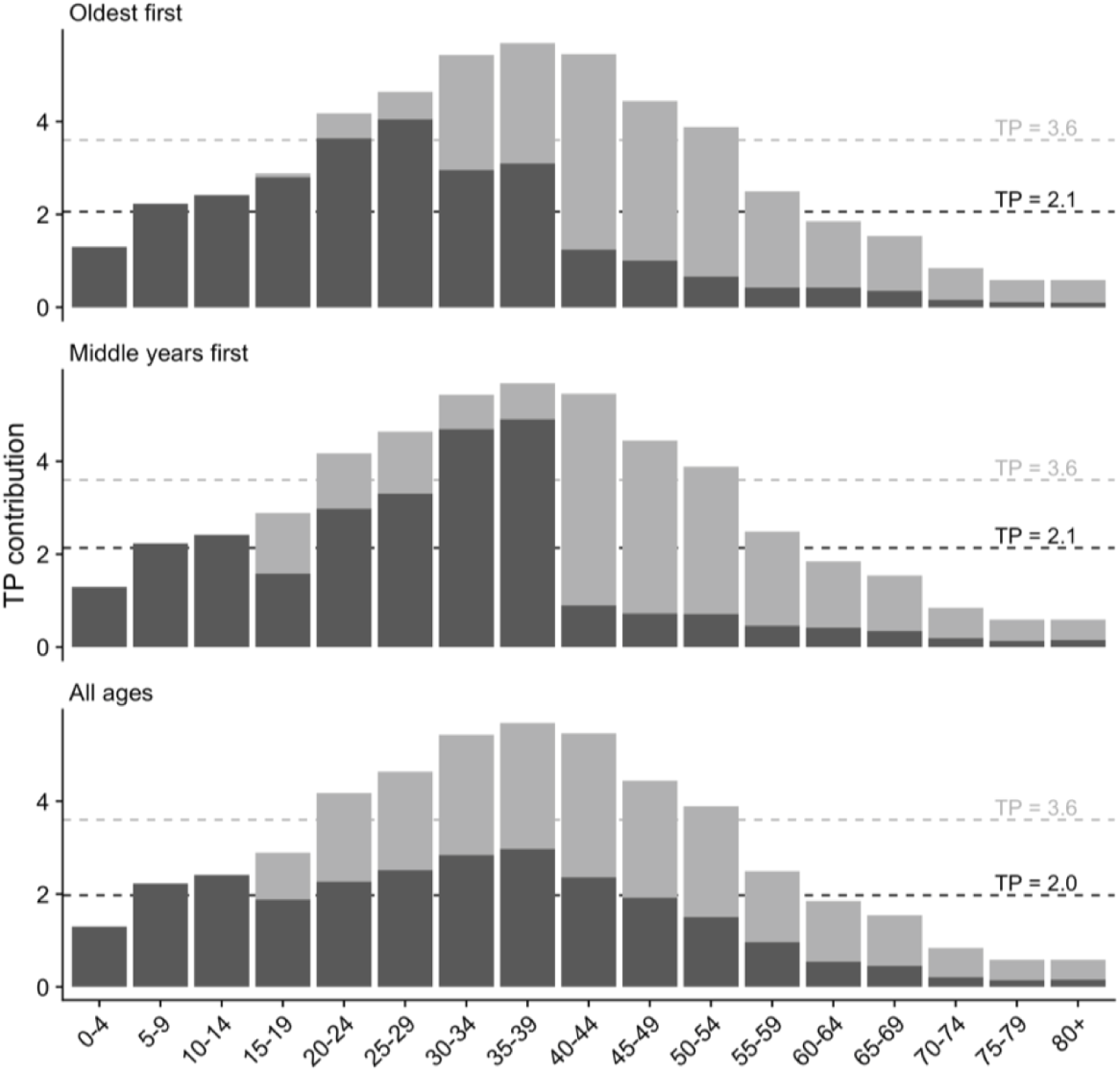
Impact of the four different vaccine allocation strategies on TP by age category, resulting in the overall TP achieved by 50% age eligible population coverage. Dark grey represents TP contribution after vaccination, and light grey in the absence of vaccination. Dashed lines correspond to whole population differences in TP with age-group interactions included (is marginal contribution of each age-group / column sums of transmission matrix)

**Figure S4.**
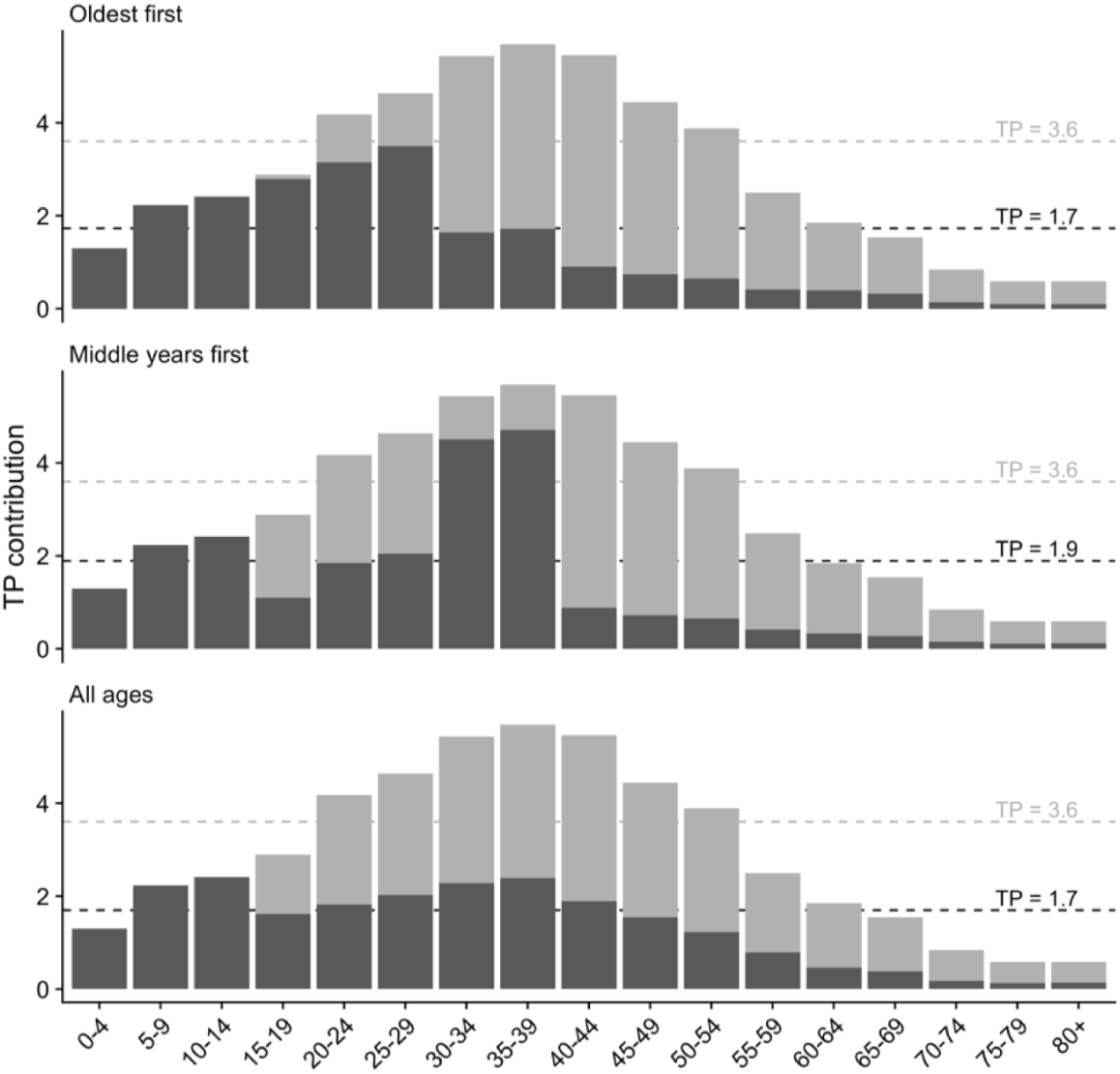
Impact of the four different vaccine allocation strategies on TP by age category, resulting in the overall TP achieved by 60% age eligible population coverage. Dark grey represents TP contribution after vaccination, and light grey in the absence of vaccination. Other coverage levels (50, 60 and 80% are in figures S2a-c). Dashed lines correspond to whole population differences in TP with age-group interactions included (is marginal contribution of each age-group / column sums of transmission matrix)

**Figure S5.**
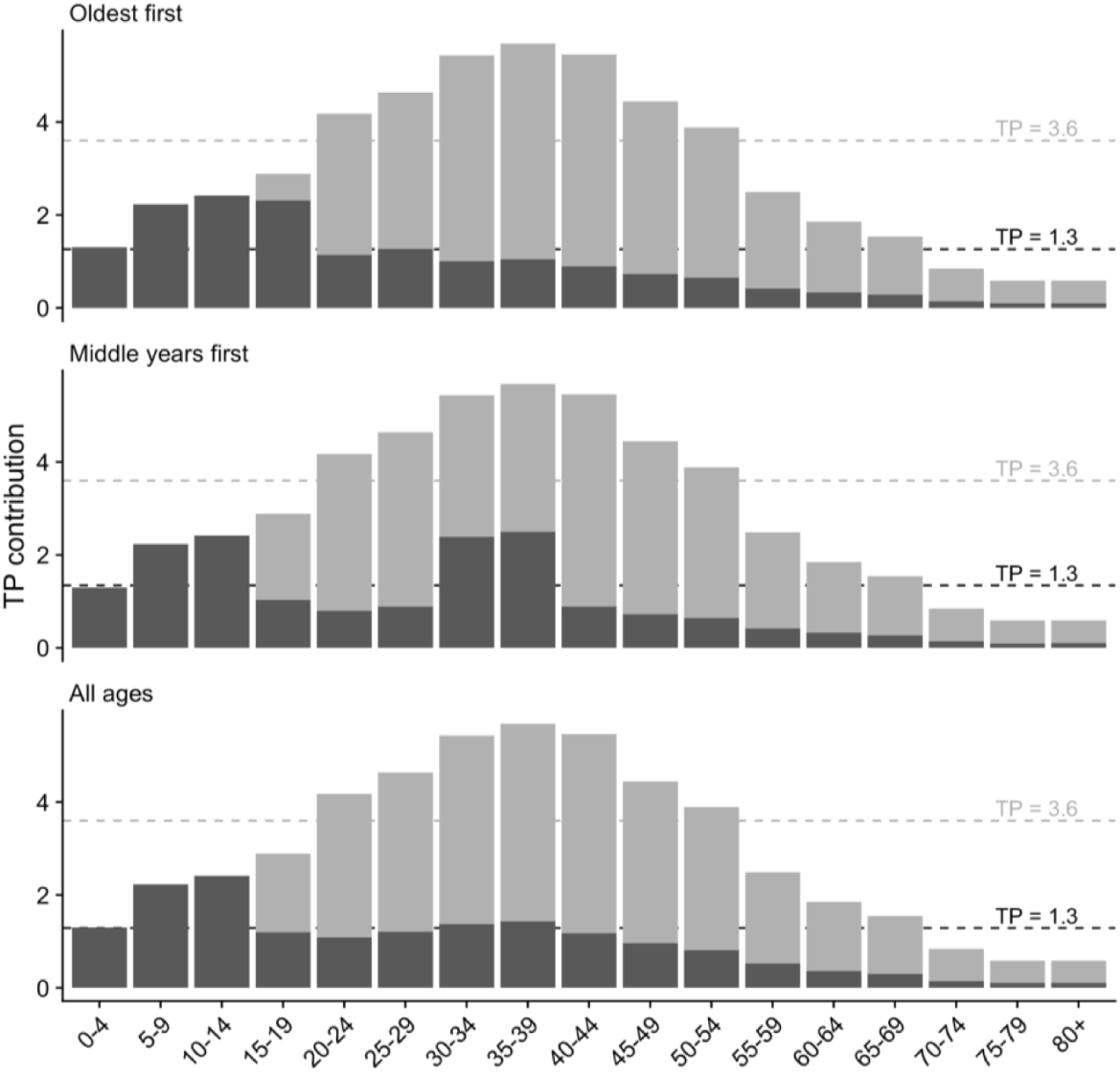
Impact of the four different vaccine allocation strategies on TP by age category, resulting in the overall TP achieved by 80% age eligible population coverage. Dark grey represents TP contribution after vaccination, and light grey in the absence of vaccination. Dashed lines correspond to whole population differences in TP with age-group interactions included (is marginal contribution of each age-group / column sums of transmission matrix)

**Table S2.** Proportion of time lockdowns are needed to constrain transmission when the TTIQ public health response is only partially effective, due to high caseloads

| Vaccine coverage | Allocation Scenario | Light restrictions only | Moderate lockdowns only | Strict lockdowns only |
| --- | --- | --- | --- | --- |
| 50% | Oldest first | Not possible to constrain outbreak | Not possible to constrain outbreak | 89% |
|  | 40+ years first | Not possible to constrain outbreak | Not possible to constrain outbreak | 93% |
|  | All adults | Not possible to constrain outbreak | Not possible to constrain outbreak | 84% |
| 60% | Oldest first | Not possible to constrain outbreak | Not possible to constrain outbreak | 67% |
|  | 40+ years first | Not possible to constrain outbreak | Not possible to constrain outbreak | 78% |
|  | All adults | Not possible to constrain outbreak | Not possible to constrain outbreak | 65% |
| 70% | Oldest first | Not possible to constrain outbreak | 77% | 47% |
|  | 40+ years first | Not possible to constrain outbreak | 99% | 60% |
|  | All adults | Not possible to constrain outbreak | 81% | 49% |
| 80% | Oldest first | 82% | 47% | 29% |
|  | 40+ years first | Not possible to constrain outbreak | 59% | 36% |
|  | All adults | 89% | 51% | 31% |

**Table S3.** Proportion of time lockdowns are needed to constrain transmission when the TTIQ public health response is optimally effective.

| Vaccine coverage | Allocation Scenario | Light restrictions only | Moderate lockdowns only | Strict lockdowns only |
| --- | --- | --- | --- | --- |
| 50% | Oldest first | Not possible to constrain outbreak | Not possible to constrain outbreak | 63% |
|  | 40+ years first | Not possible to constrain outbreak | Not possible to constrain outbreak | 67% |
|  | All adults | Not possible to constrain outbreak | 94% | 58% |
| 60% | Oldest first | Not possible to constrain outbreak | 67% | 41% |
|  | 40+ years first | Not possible to constrain outbreak | 86% | 52% |
|  | All adults | Not possible to constrain outbreak | 64% | 39% |
| 70% | Oldest first | 60% | 34% | 21% |
|  | 40+ years first | 97% | 56% | 34% |
|  | All adults | 67% | 38% | 23% |
| 80% | Oldest first | 7% | 4% | 3% |
|  | 40+ years first | 29% | 17% | 10% |
|  | All adults | 15% | 8% | 5% |

**Table S4.** Proportion of time lockdowns needed to constrain transmission when the TTIQ public health response is only partially effective, due to high caseloads, and where light restrictions are always in place.

| Vaccine coverage | Allocation Scenario | Moderate lockdowns only | Strict lockdowns only |
| --- | --- | --- | --- |
| 50% | Oldest first | Not possible to constrain outbreak | 82% |
|  | 40+ years first | Not possible to constrain outbreak | 89% |
|  | All adults | Not possible to constrain outbreak | 75% |
| 60% | Oldest first | Not possible to constrain outbreak | 49% |
|  | 40+ years first | Not possible to constrain outbreak | 67% |
|  | All adults | Not possible to constrain outbreak | 46% |
| 70% | Oldest first | 46% | 18% |
|  | 40+ years first | 97% | 39% |
|  | All adults | 55% | 22% |
| 80% | Oldest first | 0% | 0% |
|  | 40+ years first | 4% | 2% |
|  | All adults | 0% | 0% |

**Table S5.** Proportion of time lockdowns needed to constrain transmission when the TTIQ public health response is optimally effective and where light restrictions are always in place.

| Vaccine coverage | Allocation Scenario | Moderate lockdowns only | Strict lockdowns only |
| --- | --- | --- | --- |
| 50% | Oldest first | Not possible to constrain outbreak | 42% |
|  | 40+ years first | Not possible to constrain outbreak | 49% |
|  | All adults | 87% | 35% |
| 60% | Oldest first | 23% | 9% |
|  | 40+ years first | 66% | 27% |
|  | All adults | 15% | 6% |
| 70% | Oldest first | 0% | 0% |
|  | 40+ years first | 0% | 0% |
|  | All adults | 0% | 0% |
| 80% | Oldest first | 0% | 0% |
|  | 40+ years first | 0% | 0% |
|  | All adults | 0% | 0% |

## REFERENCES

1. World Health Organisation. Weekly epidemiological update on COVID-19 - 10 August 2022. Technical report, August 2022. URL https://www.who.int/publications/m/item/weekly-epidemiological-update-on-covid-1910-august-2022.

2. Ilan Noy, Nguyen Doan, and Tauisi Taupo. The economic risk from COVID-19 in Pacific Island countries: very few infections but lots of pain. New Zealand Economic Papers, 56 (1):55–66, January 2022. ISSN 0077-9954. doi: 10.1080/00779954.2020.1827016. URL https://doi.org/10.1080/00779954.2020.1827016. Publisher: Routledge eprint: https://doi.org/10.1080/00779954.2020.1827016.

3. Emeline Han, Melisa Mei Jin Tan, Eva Turk, Devi Sridhar, Gabriel M Leung, Kenji Shibuya, Nima Asgari, Juhwan Oh, Alberto L García-Basteiro, Johanna Hanefeld, Alex R Cook, Li Yang Hsu, Yik Ying Teo, David Heymann, Helen Clark, Martin McKee, and Helena Legido-Quigley. Lessons learnt from easing COVID-19 restrictions: an analysis of countries and regions in Asia Pacific and Europe. The Lancet, 396(10261):1525–1534, November 2020. ISSN 0140-6736. doi: 10.1016/S0140-6736(20)32007-9. URL https://www.sciencedirect.com/science/article/pii/S0140673620320079.

4. Kathina Ali, Matthew Iasiello, Joep van Agteren, Teri Mavrangelos, Michael Kyrios, and Daniel B. Fassnacht. A cross-sectional investigation of the mental health and wellbeing among individuals who have been negatively impacted by the COVID-19 international border closure in Australia. Globalization and Health, 18(1):12, February 2022. ISSN 1744-8603. doi: 10.1186/s12992-022-00807-7. URL https://doi.org/10.1186/s12992-022-00807-7.

5. Tung Le, Zacharias Andreadakis, Arun Kumar, Raúl Román, Stig Tollefsen, Melanie Saville, and Stephen Mayhew. The COVID-19 vaccine development landscape. Nature Reviews Drug Discovery, April 2020. doi: 10.1038/d41573-020-00073-5.

6. Alan D. T. Barrett, Richard W. Titball, Paul A. MacAry, Richard E. Rupp, Veronika von Messling, David H. Walker, and Nicolas V. J. Fanget. The rapid progress in COVID vaccine development and implementation. npj Vaccines, 7(1):1–2, February 2022. ISSN 2059-0105. doi: 10.1038/s41541-022-00442-8. URL https://www.nature.com/articles/s41541-022-00442-8. Number: 1 Publisher: Nature Publishing Group.

7. Seyed M. Moghadas, Pratha Sah, Thomas N. Vilches, and Alison P. Galvani. Can the USA return to pre-COVID-19 normal by July 4? The Lancet Infectious Diseases, 21(8):1073– 1074, August 2021. ISSN 1473-3099, 1474-4457. doi: 10.1016/S1473-3099(21)00324-8. URL https://www.thelancet.com/journals/laninf/article/PIIS1473-3099(21)00324-8/fulltext. Publisher: Elsevier.

8. Grace Ho. Zero-Covid strategy no longer feasible due to highly infectious Delta variant: PM Lee. The Straits Times, October 2021. ISSN 0585-3923. URL https://www.straitstimes.com/singapore/politics/spore-must-press-on-with-strategy-of-living-with-covid-19-and-not-be-paralysed-by.

9. Alicia Blair, Mattia de Pasquale, Valentin Gabeff, Mélanie Rufi, and Antoine Flahault. The End of the Elimination Strategy: Decisive Factors towards Sustainable Management of COVID-19 in New Zealand. Epidemiologia, 3(1):135–147, March 2022. ISSN 2673-3986. doi: 10.3390/epidemiologia3010011. URL https://www.mdpi.com/2673-3986/3/1/11. Number: 1 Publisher: Multidisciplinary Digital Publishing Institute.

10. Nick Golding, David J Price, Gerard Ryan, Jodie McVernon, James M McCaw, and Freya M Shearer. A modelling approach to estimate the transmissibility of SARS-CoV-2 during periods of high, low, and zero case incidence. eLife, 12:e78089, January 2023. ISSN 2050-084X. doi: 10.7554/eLife.78089. URL https://doi.org/10.7554/eLife.78089. Publisher: eLife Sciences Publications, Ltd.

11. Freya M. Shearer, James M. McCaw, Gerard Ryan, Tianxiao Hao, Nicholas J. Tierney, Michael Lydeamore, Kate Ward, Sally Ellis, James Wood, Jodie McVernon, and Nick Golding. Estimating the impact of test-trace-isolate-quarantine systems on SARS-CoV-2 transmission in Australia, January 2023. URL https://www.medrxiv.org/content/10.1101/2023.01. 10.23284209v1. Pages: 2023.01.10.23284209.

12. Australian Government. National Plan to transition Australia’s National COVID-19 Response. Technical report, 2021. URL https://pmtranscripts.pmc.gov.au/sites/default/files/2022-06/national-plan-060821_0.pdf. natplan.

13. Department of the Treasury. National Plan to Transition to Australia’s National COVID-19 Response: Economic Impact Analysis. Technical report, 2021. URL https://treasury.gov.au/sites/default/files/2021-08/PDF_Economic_Impacts_COVID-19_Response_196731.pdf.

14. Doherty Modelling Group. DOHERTY MODELLING –FINAL REPORT TO NATIONAL CABI-NET. Technical report, November 2021. URL https://www.doherty.edu.au/uploads/content_doc/Synthesis_DohertyModelling_FinalReport__NatCab05Nov.pdf.

15. Doherty Modelling Group. DOHERTY MODELLING INTERIM REPORT TO NATIONAL CABINET. Technical report, September 2021. URL https://www.doherty.edu.au/uploads/content_doc/DOHERTY_MODELLING_INTERIM_REPORT_TO_NATIONAL_CABINET_17TH_SEPTEMBER_2021.pdf.

16. Doherty Modelling Group. DOHERTY MODELLING REPORT REVISED. Technical report, August 2021. URL http://www.doherty.edu.au/uploads/content_doc/DohertyModelling_NationalPlan_and_Addendum_20210810.pdf.

17. Department of Health and Aged Care. Coronavirus (COVID-19) Common Operating Picture – 24 May 2022. Technical report, May 2022. URL https://www.health.gov.au/resources/publications/coronavirus-covid-19-common-operating-picture-24-may-2022.

18. Nick Golding, Freya M Shearer, Robert Moss, Peter Dawson, Dennis Liu, Joshua V Ross, Rob Hyndman, Pablo Montero-Manso, Gerry Ryan, Tobin South, Jodie McVernon, David J Price, and James M McCaw. Situational assessment of COVID-19 in Australia Technical Report 15 March 2021 (released 28 May 2021). Technical report, May 2021.

19. James M McCaw, Robert Moss, David J Price, Freya M Shearer, Nick Golding, Tianxiao Hao, Gerry Ryan, Adeshina Adekunle, Peter Dawson, Mingmei Teo, Dylan Morris, Joshua V Ross, Tobin South, Rob Hyndman, Michael Lydeamore, Mitchell O’Hara-Wild, and James Wood. Situational assessment of COVID-19 in Australia Technical Report 22 May 2022 (released 12 August 2022). Technical report, August 2022. URL https://mspgh.unimelb.edu.au/data/assets/pdf_file/0003/4256103/2022-05-22-Technical-report-public-release.pdf.

20. Don Klinkenberg, Christophe Fraser, and Hans Heesterbeek. The Effectiveness of Contact Tracing in Emerging Epidemics. PLOS ONE, 1(1):e12, December 2006. ISSN 1932-6203. doi: 10.1371/journal.pone.0000012. URL https://journals.plos.org/plosone/article?id=10.1371/journal.pone.0000012. Publisher: Public Library of Science.

21. Lawrence O. Gostin, Ronald Bayer, and Amy L. Fairchild. Ethical and Legal Challenges Posed by Severe Acute Respiratory Syndrome: Implications for the Control of Severe Infectious Disease Threats. JAMA, 290(24):3229–3237, December 2003. ISSN 0098-7484. doi: 10.1001/jama.290.24.3229. URL https://doi.org/10.1001/jama.290.24.3229.

22. Peter Ashcroft, Sonja Lehtinen, and Sebastian Bonhoeffer. Test-trace-isolate-quarantine (TTIQ) intervention strategies after symptomatic COVID-19 case identification. PLOS ONE, 17(2):e0263597, February 2022. ISSN 1932-6203. doi: 10.1371/journal.pone.0263597. URL https://journals.plos.org/plosone/article?id=10.1371/journal.pone.0263597. Publisher: Public Library of Science.

23. Marcel Salathé, Christian L. Althaus, Richard Neher, Silvia Stringhini, Emma Hodcroft, Jacques Fellay, Marcel Zwahlen, Gabriela Senti, Manuel Battegay, Annelies Wilder-Smith, Isabella Eckerle, Matthias Egger, and Nicola Low. COVID-19 epidemic in Switzerland: on the importance of testing, contact tracing and isolation. Swiss Medical Weekly, (11), March 2020. doi: 10.4414/smw.2020.20225. URL https://smw.ch/article/doi/smw.2020.20225. Publisher: EMH Media.

24. World Health Organisation. WHO SAGE roadmap for prioritizing uses of COVID-19 vaccines in the context of limited supply: an approach to inform planning and subsequent recommendations based on epidemiological setting and vaccine supply scenarios, first issued 20 October 2020, latest update 16 July 2021. Technical report, World Health Organization, July 2021. URL https://apps.who.int/iris/handle/10665/342917.

25. Department of Health and Aged Care. Op COVID SHIELD National COVID Vaccine Campaign Plan. Technical report, August 2021. URL https://www.health.gov.au/resources/publications/op-covid-shield-national-covid-vaccine-campaign-plan.

26. Therapeutic Goods Administration. AUSTRALIAN PRODUCT INFORMATION – COMIRNATY® (tozinameran) COVID-19 VACCINE [Tris/Sucrose Presentation], 2022. URL https://www.tga.gov.au/covid-19-vaccine-pfizer-australia-comirnaty-tozinameran-mrna#documents.

27. Therapeutic Goods Administration. AUSTRALIAN PRODUCT INFORMATION VAXZEVRIA® (previously COVID-19 Vaccine AstraZeneca) (ChAdOx1-S) solution for injection, 2022. URL https://www.tga.gov.au/covid-19-vaccine-astrazeneca-vaxzevria.

28. Australian Technical Advisory Group on Immunisation. ATAGI statement on revised recommendations on the use of COVID-19 Vaccine AstraZeneca, 17 June 2021, June 2021. URL https://www.health.gov.au/news/atagi-statement-on-revised-recommendations-on-the-use-of-covid-19-vaccine-astrazeneca-17-june-2021. Publisher: Australian Government Department of Health and Aged Care.

29. Kiesha Prem, Alex R. Cook, and Mark Jit. Projecting social contact matrices in 152 countries using contact surveys and demographic data. PLOS Computational Biology, 13(9):e1005697, September 2017. ISSN 1553-7358. doi: 10.1371/journal.pcbi.1005697. URL https://journals.plos.org/ploscompbiol/article?id=10.1371/journal.pcbi.1005697. Publisher: Public Library of Science.

30. Nicholas G. Davies, Petra Klepac, Yang Liu, Kiesha Prem, Mark Jit, and Rosalind M. Eggo. Age-dependent effects in the transmission and control of COVID-19 epidemics. Nature Medicine, 26(8):1205–1211, August 2020. ISSN 1546-170X. doi: 10.1038/s41591-020-0962-9. URL https://www.nature.com/articles/s41591-020-0962-9. Number: 8 Publisher: Nature Publishing Group.

31. R Core Team. R: A Language and Environment for Statistical Computing. R Foundation for Statistical Computing, Vienna, Austria, 2021. URL https://www.R-project.org.

32. Gerard Ryan and Nick Golding. geryan/nat plan figs: Citable version for national plan paper, September 2022. URL 10.5281/zenodo.7117840.

33. Alexandra B. Hogan, Peter Winskill, Oliver J. Watson, Patrick G. T. Walker, Charles Whittaker, Marc Baguelin, Nicholas F. Brazeau, Giovanni D. Charles, Katy A. M. Gaythorpe, Arran Hamlet, Edward Knock, Daniel J. Laydon, John A. Lees, Alessandra Løchen, Robert Verity, Lilith K. Whittles, Farzana Muhib, Katharina Hauck, Neil M. Ferguson, and Azra C. Ghani. Within-country age-based prioritisation, global allocation, and public health impact of a vaccine against SARS-CoV-2: A mathematical modelling analysis. Vaccine, 39(22):2995–3006, May 2021. ISSN 0264-410X. doi: 10.1016/j.vaccine.2021.04.002. URL https://www.sciencedirect.com/science/article/pii/S0264410X21004278.

34. Gareth Iacobucci. Covid-19: “Freedom day” in England could lead to “significant third wave of hospitalisations and deaths,” modelling predicts. BMJ, 374:n1789, July 2021. ISSN 1756-1833. doi: 10.1136/bmj.n1789. URL https://www.bmj.com/content/374/bmj.n1789. Publisher: British Medical Journal Publishing Group Section: News.

35. Philip Ball. Why England’s COVID ‘freedom day’ alarms researchers. Nature, 595(7868):479–480, July 2021. doi: 10.1038/d41586-021-01938-4. URL https://www.nature.com/articles/d41586-021-01938-4. Bandiera abtest: a Cg type: News Number: 7868 Publisher: Nature Publishing Group Subject term: Epidemiology, Virology.

36. OECD. First lessons from government evaluations of COVID-19 responses: A synthesis. Technical report, Organisation for Economic Co-operation and Development, January 2022. URL https://www.oecd.org/coronavirus/policy-responses/first-lessons-from-government-evaluations-of-covid-19-responses-a-synthesis-483507d6/.

37. Yinon M. Bar-On, Yair Goldberg, Micha Mandel, Omri Bodenheimer, Laurence Freedman, Nir Kalkstein, Barak Mizrahi, Sharon Alroy-Preis, Nachman Ash, Ron Milo, and Amit Huppert. Protection of BNT162b2 Vaccine Booster against Covid-19 in Israel. New England Journal of Medicine, 385(15):1393–1400, October 2021. ISSN 0028-4793. doi: 10.1056/NEJMoa2114255. URL https://doi.org/10.1056/NEJMoa2114255. Publisher: Massachusetts Medical Society eprint: https://doi.org/10.1056/NEJMoa2114255.

38. The Lancet Infectious Diseases. COVID-19 vaccine equity and booster doses. The Lancet Infectious Diseases, 21(9):1193, September 2021. ISSN 1473-3099, 1474-4457. doi: 10.1016/S1473-3099(21)00486-2. URL https://www.thelancet.com/journals/laninf/article/PIIS1473-3099(21)00486-2/fulltext#coronavirus-linkback-header. Publisher: Elsevier.

39. Eamon Conway, Camelia Walker, Chris Baker, Michael Lydeamore, Gerard E. Ryan, Trish Campbell, Joel C. Miller, Max Yeung, Greg Kabashima, James Wood, Nic Rebuli, James M. McCaw, Jodie McVernon, Nick Golding, David J. Price, and Freya M. Shearer. COVID-19 vaccine coverage targets to inform reopening plans in a low incidence setting, December 2022. URL https://www.medrxiv.org/content/10.1101/2022.12.04.22282996v1. Pages: 2022.12.04.22282996.

40. David S Khoury, Deborah Cromer, Arnold Reynaldi, Timothy E Schlub, Adam K Wheatley, Jennifer A Juno, Kanta Subbarao, Stephen J Kent, James A Triccas, and Miles P Davenport. Neutralizing antibody levels are highly predictive of immune protection from symptomatic sars-cov-2 infection. Nature medicine, 27(7):1205–1211, 2021.

